# A novel loop-mediated isothermal amplification (LAMP) primer set for detecting the STY2879 gene of *Salmonella enterica* serovar Typhi in raw milk

**DOI:** 10.1101/2025.03.26.25324623

**Authors:** Hyuck-Jin Seo, Timothy E. Riedel

## Abstract

Milk-transmitted outbreaks remain a significant issue in low-income countries due to unhygienic practices. Currently, there are no known easily accessible and cheap diagnostic tests that can detect *Salmonella enterica* subsp. e*nterica* serovar Typhi, the causative agent of typhoid fever, in raw milk samples at high specificity, sensitivity, and speed without preprocessing. Using colorimetric loop-mediated isothermal amplification, we screened 15 novel and two previously published primer sets. We identified one novel primer set capable of detecting the STY2879 gene in as few as 2000 genomes of *Salmonella enterica* subsp. e*nterica* serovar Typhi in 2% diluted raw milk samples after a 30-minute incubation at 65° C. The colorimetric readout offers potential applications for on-site detection in remote and low-resource settings where *Salmonella enterica* subsp. e*nterica* serovar Typhi remains a public health concern.

## 1. Introduction

*Salmonella enterica* subsp. e*nterica* serovar Typhi (*S*. Typhi) is a highly pathogenic, human-restricted bacterium responsible for typhoid fever, a systemic illness characterized by fever, headache, malaise, abdominal pain, and, in severe cases, death. Globally, in 2017, *S*. Typhi infected approximately 10.9 million people and resulted in nearly 120,000 deaths [1]. Transmission typically occurs through the ingestion of contaminated food or water, and unpasteurized raw milk has increasingly been recognized as a vehicle of transmission. A recent study from Ecuador analyzed 600 raw milk samples and reported a 37.5% prevalence of *Salmonella enterica* contamination. Among the isolates, 28 were identified as *S*. Typhi, and 19.1% of the *Salmonella*-positive samples could not be assigned a specific serovar, suggesting that the true prevalence of *S*. Typhi contamination may be even higher than reported in raw milk [2].

In developed and high-income countries (HICs), the burden of global milk-borne outbreaks is low, as nearly all milk is pasteurized and maintained under food safety standards [3]. However, in low-income countries (LICs) such as Kenya, Ethiopia, and other East African countries, the burden of milk-borne outbreaks remains high. Farmers were found to lack adequate training in milk safety standards, and milking conditions were unhygienic as the majority did not engage in teat dipping, used plastic containers, and used untreated water [4]. Furthermore, pastoral communities in such LICs were strongly opposed to participating in pasteurization practices due to a misconception that pasteurization would deplete the milk of its nutrients. As a result, pastoralists were largely found to consume unpasteurized raw milk from markets and directly from animal udders [5].

Currently, there are no known readily available diagnostic tests that can accurately detect *S*. Typhi in raw milk samples with the high specificity, sensitivity, speed, and low cost needed to be effective in LICs. Loop-mediated isothermal amplification (LAMP) is a broadly adopted nucleic acid amplification system that has the potential to overcome such shortcomings [6,7]. While similar to PCR, LAMP reactions do not require thermocyclers and can be carried out with a simple constant temperature water bath or heater device [8]. LAMP reactions utilize four to six primers, allowing for rapid amplification while maintaining a high level of specificity. This makes LAMP a strong candidate for diagnostic applications in developing and LICs with limited resources [9]. In addition, ready-to-use colorimetric LAMP systems are available that allow for easy end-user color-change interpretation. Colorimetric LAMP assays sometimes have issues with non-specific amplification products developing during the incubation period, which can lead to false positives [10]. This problem can be mitigated with additional error-checking tools, such as Jiang et al. [11], but these often lead to increased complexity of the diagnostic systems and reduce the low-resource advantage.

In this study, we aimed to develop an improved, field-deployable LAMP assay for the detection of *S*. Typhi in raw milk. To accomplish this, we systematically evaluated existing and newly designed primer sets for delayed non-specific amplification, a critical determinant of assay reliability in colorimetric LAMP systems. We screened 15 novel primer sets and two previously published sets using the NEB colorimetric LAMP system and quantified their performance based on time to amplification. Primer sets exhibiting the longest delays in non-specific amplification were further evaluated by measuring the time differential between positive and negative control amplification (defined in this study as differential time to threshold or DTT). A larger DTT indicates a lower risk of false-positive interpretation. The primer set with the highest DTT was subsequently assessed for robustness at lower signal concentrations and with the direct addition of milk.

## 2. Materials and Methods

### 2.1. Primer Sets and Positive Control Sequence

We tested two previously reported LAMP primer sets for the detection of *Salmonella* Typhi (Table 1). We also designed 15 novel primer sets specific to a validated [12–14] segment within the STY2879 target gene (Figure 1) using the NEB LAMP Primer Design Tool Version 1.4.2 [15]. Two base pairs were added to the segment to improve the likelihood of loop primer design (Table S1). LAMP primers were designed following NEB-recommended parameters (Table 2) optimized for assay robustness, specificity, and reaction speed in the NEB colorimetric LAMP system at a reaction incubation temperature of 65 °C [16].

**Table 1.**
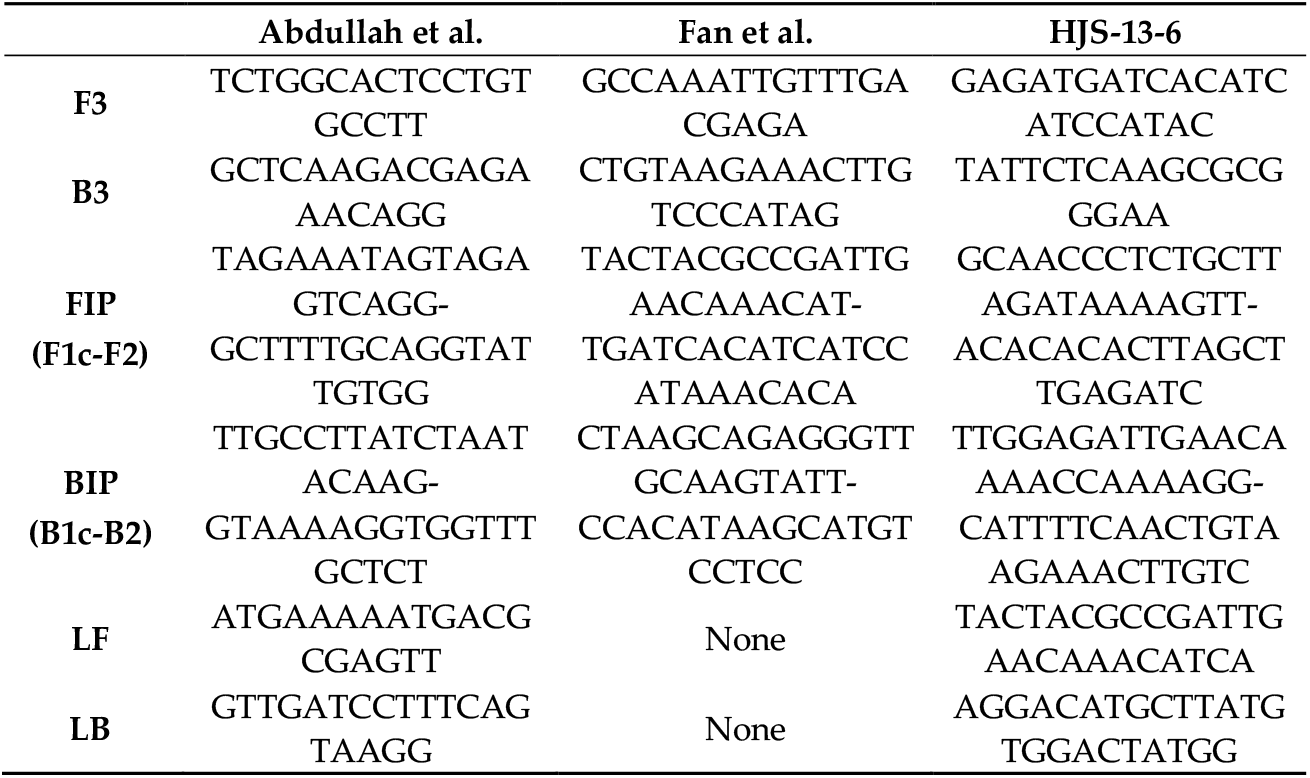
Loop-mediated isothermal amplification (LAMP) primer sets screened for their performance in the NEB colorimetric LAMP system, shown in the 5’ to 3’ direction. Abdullah et al. [17] primers targeted the STBHUCCB_38510 gene, while Fan et al. [12] and HJS-13-6 (this study) primers both targeted the STY2879 gene. This study developed 15 new primer sets for the STY2879 gene. Primer set HJS-13-6, shown here, exhibited the longest incubation time before negative controls showed amplification. The remaining 14 primer sets are shown in Table S2.

**Table 2.** LAMP primer design parameters used to generate 15 novel primer sets evaluated in this study. These parameters enforce optimal amplicon length, loop primer melting temperature, controlled GC content, terminal stability, and minimized secondary structure formation to maximize assay speed, sensitivity, and specificity in the NEB colorimetric LAMP system at a reaction incubation temperature of 65 °C.

| Category | Parameter | Value / Range |
| --- | --- | --- |
| Reaction Conditions (mM) | Na<sup>+</sup> concentration | 50 |
|  | Mg<sup>2+</sup> concentration | 8 |
| Primer Length (bp) | F1c/B1c | 20-22 |
|  | F2/B2 | 18-20 |
|  | F3/B3 | 18-20 |
|  | LF/LB | 15-25 |
| Melting Temperature (°C) | F1c/B1c | 64-66 |
|  | F2/B2 | 59-61 |
|  | F3/B3 | 59-61 |
|  | LF/LB | 64-66 |
| GC Content (%) | All primers | 40-65 |
|  | Loop primers | 40-65 |
| Terminal Stability<br>($\Delta G$, kcal·mol<sup>-1</sup>) | 5' end | ≤ -4 |
|  | 3' end | ≤ -4 |
|  | 3' end (loop primers) | ≤ -2 |
| Secondary Structure Thresholds<br>($\Delta G$, kcal·mol<sup>-1</sup>) | Dimer check | ≤ -2.5 |
|  | Dimer check (loop primers) | ≤ -3.5 |
| Inter-Primer Distances (bp) | F2-B2 (amplicon length) | 120-160 |
|  | F1c-F2 (loop region) | 40-60 |
|  | F2-F3 | 0-60 |
|  | F1c-B1c | 0-100 |
| Primer Count Limits | F1c/B1c | ≤ 3 |
|  | F2/B2 | ≤ 10 |
|  | F3/B3 | ≤ 3 |
|  | LF/LB | ≤ 10 |

**Figure 1.**
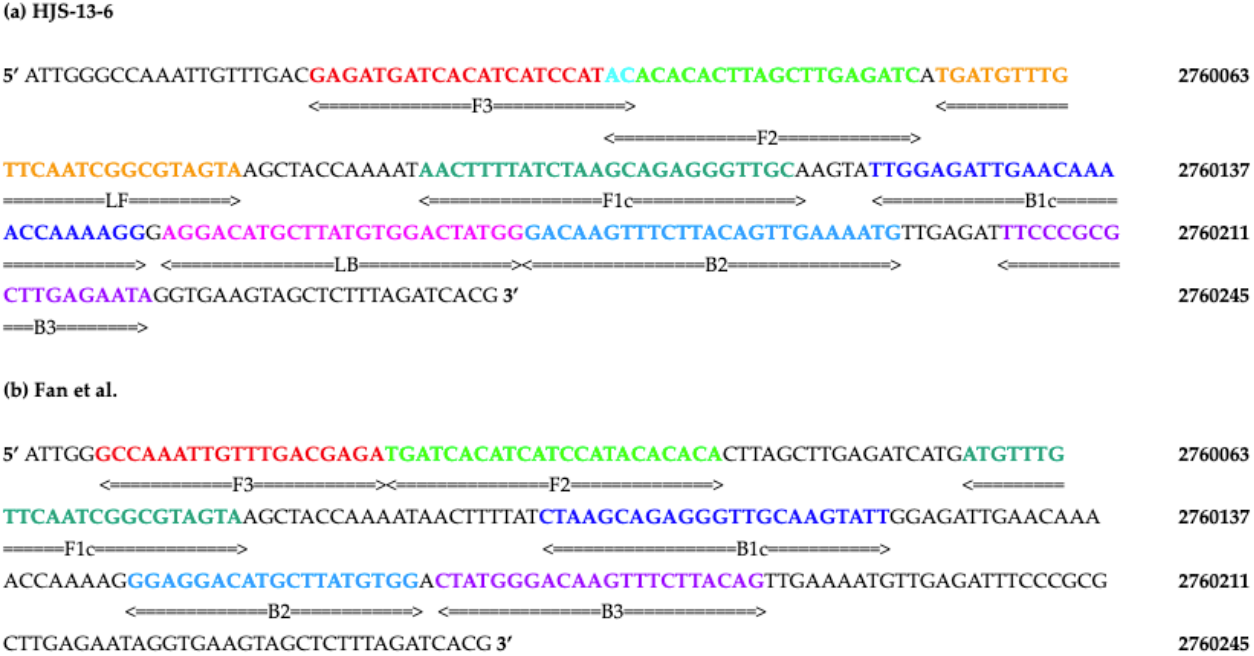
Comparison of primer binding sites between the (a) newly designed HJS-13-6 primer set and the (b) Fan et al. [12] primer set. The template sequence used for designing both sets is a segment within the STY2879 gene (shown 5’ to 3’). Colored regions represent individual primer binding sites: F3 (in red), FIP (in dark and light green), BIP (in dark and light blue), and B3C (in purple). The HJS-13-6 primer set includes a 2-base-pair overlap (in cyan) between the F3 and F2 regions that enables the NEB LAMP Primer Design Tool [15] to generate loop primers: LF (in orange) and LB (in pink). Nucleotide numbering corresponds to the *S*. Typhi strain CT-18 genome (NCBI Reference Sequence: NC_003198.1).

### 2.2. Colorimetric LAMP Assay Construction

We utilized a colorimetric LAMP system as a qualitative measure for the amplification of the target DNA sequence (New England BioLabs, Ipswich MA, USA, cat. # M1800L). All water used in this study was RNase-, DNase-, and protease-free molecular biology grade water (Corning, Manassas VA, USA; cat. #46-000-CM). All LAMP reactions described in this study, including Abdullah et al. [17] and Fan et al. [12], followed the recommended settings (temperatures and primer concentrations) outlined in the WarmStart Colorimetric LAMP 2X Master Mix Typical LAMP Protocol (details described below). These LAMP reactions change color from pink to yellow due to pH changes resulting from DNA amplification. In 0.2 mL tubes, 25 µL reactions were constructed with the following: 12.5 µL of NEB 2X Colorimetric Master Mix (contains all necessary ingredients for colorimetric LAMP except primers and template), 8 µL water, and 2.5 µL of a 10X mix of all primers (stored frozen until use) for final primer concentrations of 0.2 µM Forward Outer Primer (F3) and Backward Outer Primer (B3), 1.6 µM Forward Inner Primer (FIP, composed of F1c and F2 regions) and Backward Inner Primer (BIP, composed of B1c and B2 regions), and 0.4 µM Loop Forward (LF) and Loop Backward (LB). The above components (Table 3) bring the volume in each reaction tube to 23 µL. Two additional microliters were added before incubation and are described below.

**Table 3.** Stock and final concentrations of primers for all LAMP reactions reported in this study.

| Primer | 10X Stock ($\mu\text{M}$) | 1X Reaction ($\mu\text{M}$) |
| --- | --- | --- |
| **F3** | 2 | 0.2 |
| **B3** | 2 | 0.2 |
| **FIP** | 16 | 1.6 |
| **BIP** | 16 | 1.6 |
| **LF** | 4 | 0.4 |
| **LB** | 4 | 0.4 |

For non-template negative controls (NCs), the final two microliters were used to inoculate reactions with a sample matrix, either water, 25% V/V diluted cow’s whole milk, or 25% V/V diluted cow’s raw milk. Dilutions were performed on the milk matrices using water on the same day as the experimental runs.

For positive controls (PCs), the final two microliters of the reaction were used to inoculate reactions with synthetic double-stranded DNA (gBlock, Integrated DNA Technologies) of the STY2879 target gene fragment (Table S1). The dehydrated double-stranded synthetic DNA was originally resuspended with Tris-EDTA buffer (Fisher BioReagents, pH 8.0) to a concentration of 10^10^ copies of DNA/µL and stored frozen at -20 °C in multiple aliquots. Before each experiment, serial dilutions were performed with either water or 25% milk on a previously unused thawed aliquot of 10^10^ copies of DNA/µL to create a final concentration of either 10^5^ or 10^3^ copies of DNA/µL. In later reactions, the primer set HJS-13-6 was validated with genomic DNA isolated from *Salmonella enterica* serovar Typhi strain Ty2 (GenBank: AE014613.1; BEI Resources, NIAID, NIH: Genomic DNA from *Salmonella enterica* subsp. *enterica*, Strain Ty2 serovar Typhi, NR-543) as the final two microliters. The genomic DNA was resuspended with TE buffer to a concentration of 10^7^ genome copies/µL and stored frozen at -20 °C in multiple aliquots. Before each experiment, serial dilutions were performed with either water or 25% milk on a previously unused thawed aliquot of 10^7^ genome copies/µL to create a final concentration of 10^3^ genome copies/µL. For reactions involving milk, some were spiked with 2 µL of the 10^3^ genome copies/µL dilution for a final concentration of 2000 genome copies in a matrix of 2% milk. Other milk reactions were spiked with 1 µL of 10^3^ genome copies/µL or 1 µL of 10^3^ copies of DNA/µL for a final concentration of 1000 gene copies in a matrix of 1% milk. The 1 µL spiked reactions were supplemented with another 1 µL of water to keep the volume consistent.

### 2.3. Incubating LAMP Reactions and Scoring Results

Strips of qPCR tubes containing the reaction mixtures were inserted in a water bath float and incubated in a laboratory water bath set to 65 °C. Reactions were analyzed every 10 minutes of incubation by removing the water bath float from the incubator, photographing the reactions (Figure 2), and then returning the water bath float to the incubator, taking approximately 10 seconds. Reaction colors were scored with the unaided eye as either pink, indicating no amplification; orange, indicating intermediate levels of amplification; or yellow, indicating full amplification until all the reactions had turned yellow.

**Figure 2.**
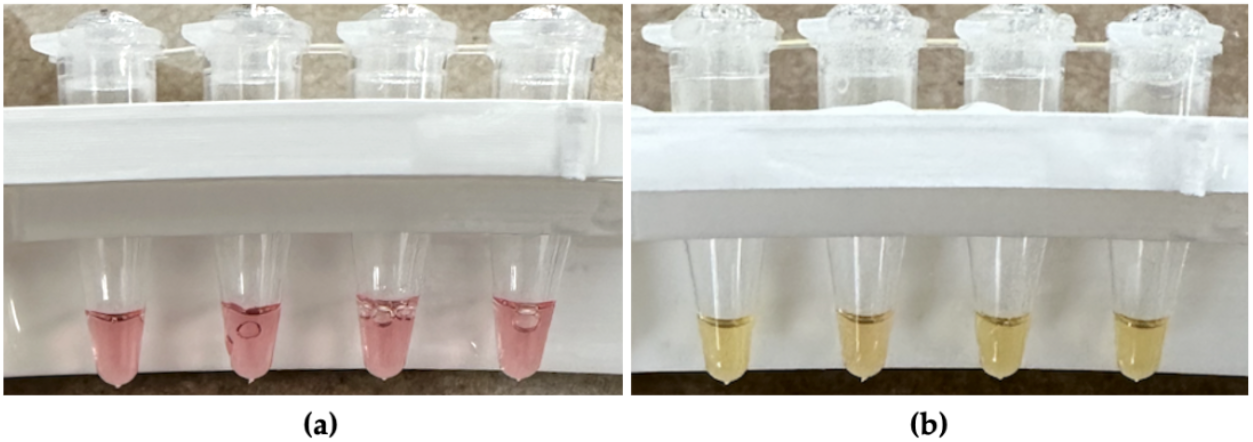
Photographs illustrating the colorimetric response of four parallel LAMP reactions during a representative screening experiment. The tubes shown are replicate reactions from a single trial evaluating the performance of primer set HJS-13-4 in a water matrix containing 10^5^ copies of DNA/µL of the STY2879 gene (Figure 3). Images were captured after temporarily removing the tubes from incubation in a 65 °C water bath at defined time points. The left panel (a) was taken after 10 minutes of incubation, when all reactions remained pink, indicating no detectable amplification. The right panel (b) was taken after 70 minutes of incubation, when all reactions had turned yellow, indicating full DNA amplification.

## 3. Results

### 3.1. Initial Screening of Primer Sets

To evaluate the performance of candidate primer sets targeting the STY2879 gene, an initial comparative screening was conducted using only NC reactions prepared in a water matrix (Figure 3). This screening was designed to assess non-specific amplification in the absence of target DNA. Primer sets that remained negative (pink) for the longest duration were considered to have the slowest non-specific amplification. The primer sets with the slowest median NC amplification time were HJS-13-6, HJS-13-3, and HJS-35. Primer set HJS-13-6 exhibited an NC amplification time of 70 minutes, and primer sets HJS-13-3 and HJS-35 exhibited an NC amplification time of 60 minutes. The three primer sets were selected for further screening.

**Figure 3.**
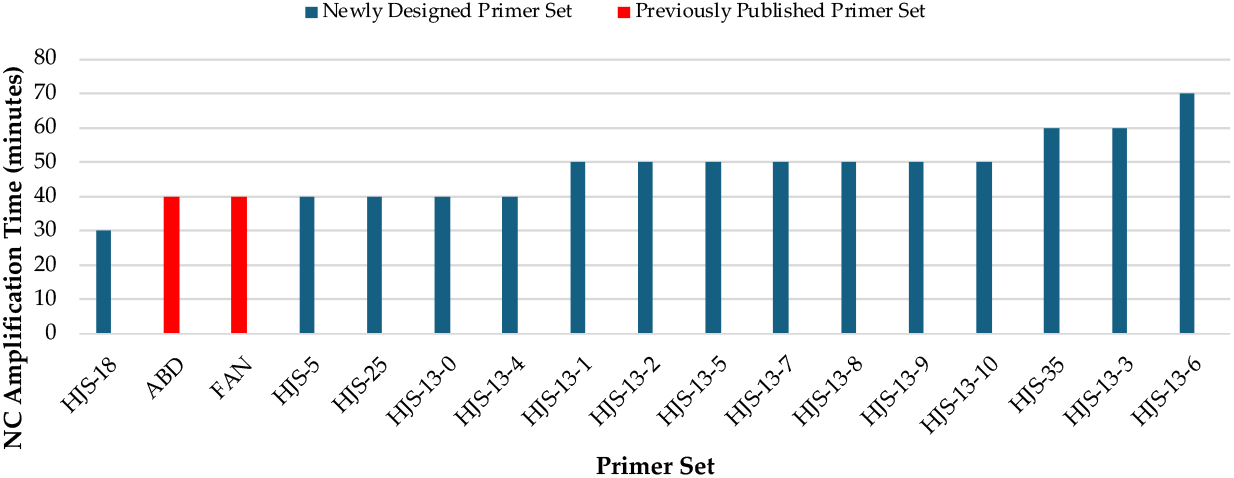
Non-template negative control (NC) amplification times. Each bar in the chart represents the median of at least 4 replicates of a primer set tested in a water matrix without positive controls (PCs). The blue bars represent newly designed primer sets (this study, Table 1 and Table S2) and the red bars represent previously published primer sets, Abdullah et al. [17] and Fan et al. [12].

Primer sets HJS-13-6, HJS-13-3, and HJS-35 were subsequently evaluated for amplification efficiency and discrimination capability using the Differential Time to Threshold (DTT) as the primary performance metric. DTT is defined in this study as the time difference between the amplification onset of the PCs and the amplification onset of the NCs. For each primer set, reactions containing a defined concentration of the STY2879 gene fragment as the PC and corresponding NC reactions were run in parallel under identical conditions. The onset time of visible color change in each reaction was recorded, and DTT values were calculated accordingly. The three primer sets were compared in a water matrix using the STY2879 gene fragment as the PC at a concentration of 10^5^ copies of DNA/µL. Primer set HJS-13-3 had a DTT of 20 minutes, primer set HJS-13-6 had the greatest DTT of 40 minutes, while primer set HJS-35 showed no ability to discriminate PCs from NCs with a DTT of 0 minutes (Figure 4).

**Figure 4.**
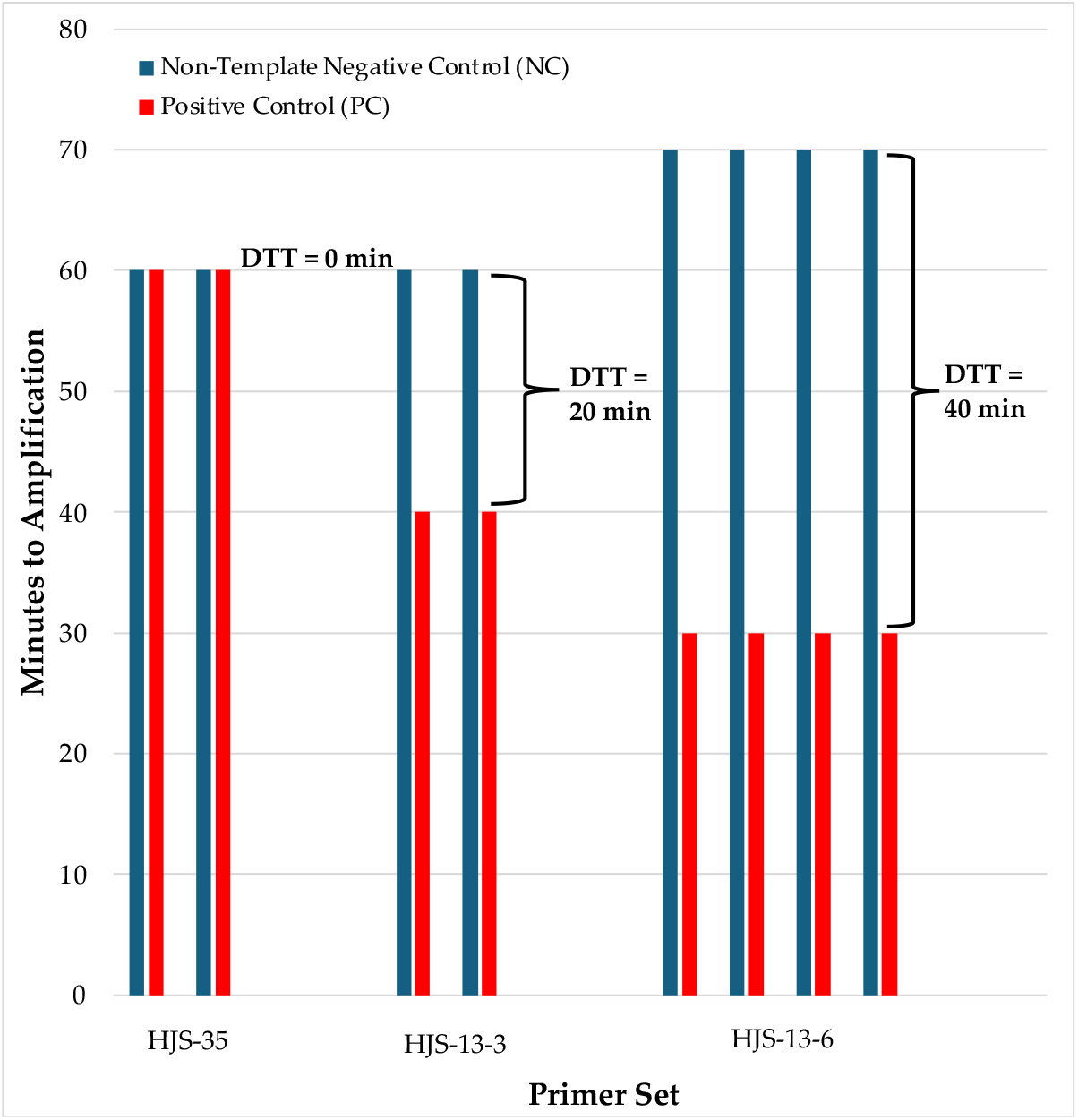
Differential Time to Thresholds (DTT) of the three longest amplifying NC primer sets. DTT is the difference in amplification onset for NC (blue bars) and PC reactions (red bars). Longer DTTs are desirable for LAMP assays. Reactions for a given primer set were tested in a water matrix containing 10^5^ copies/µL of the STY2879 gene. Braces demonstrate the DTT value between corresponding NC and PC reactions.

### 3.2. Validation of Novel Primer Set HJS-13-6

The effectiveness of primer set HJS-13-6 was further characterized in raw milk spiked with either the STY2879 gene fragment used previously or with genomic DNA isolated from *S*. Typhi strain Ty2. When tested using the STY2879 gene fragment at a concentration of 10^5^ copies of DNA/µL in a water matrix, primer set HJS-13-6 showed a DTT of 40 minutes. When spiking with a lower concentration of 10^3^ copies of DNA/µL of the STY2879 gene, a DTT of 30 minutes was observed. When spiked with 25% diluted whole milk and 10^3^ copies of DNA/µL of the STY2879 gene fragment, the DTT was 20 minutes. Changing the 25% diluted whole milk matrix to a 25% diluted raw milk matrix resulted in a DTT of 30 minutes (Table 4).

**Table 4.**
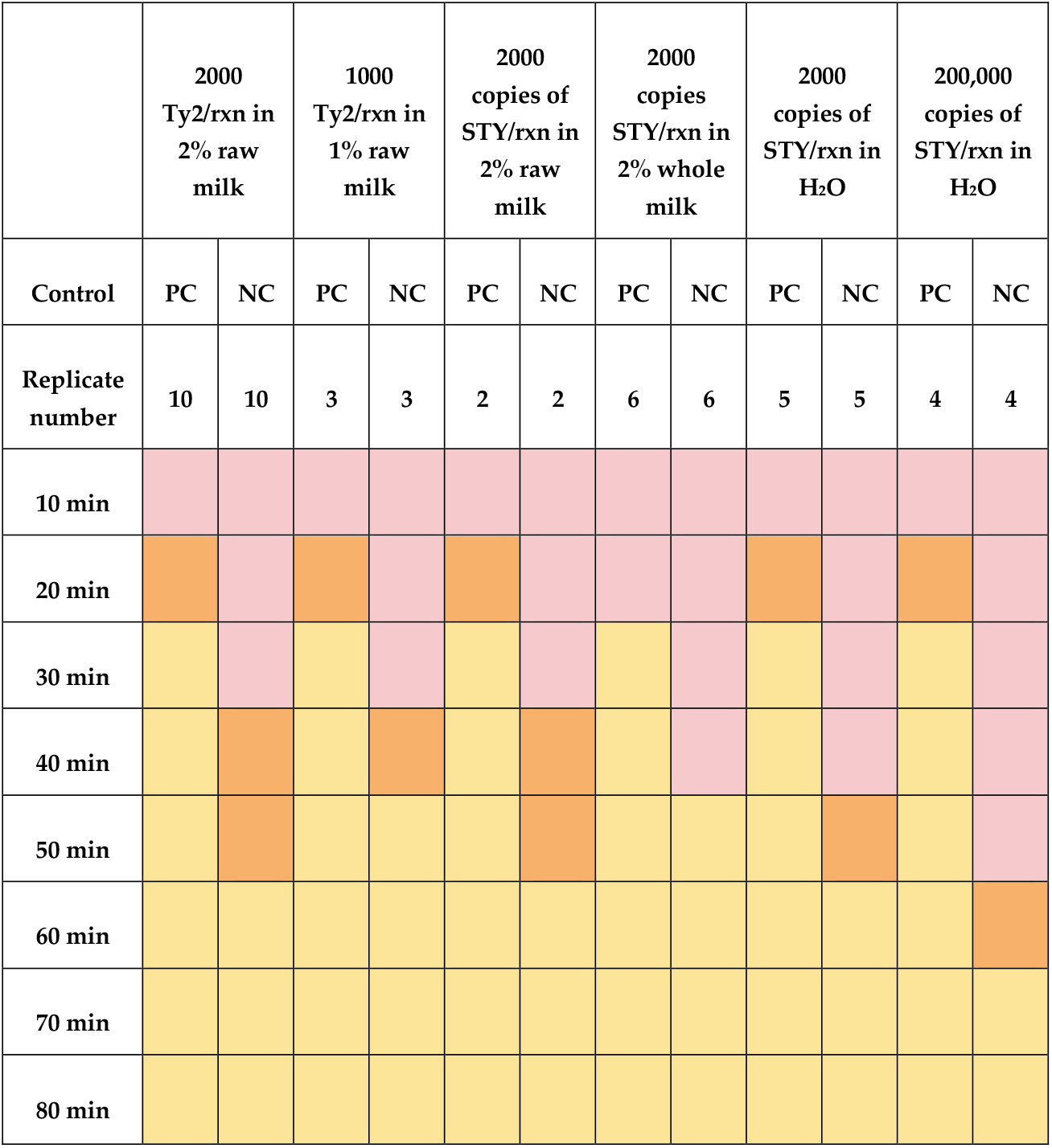
Amplification of primer set HJS-13-6 in different matrices, with different positive controls (Ty2 indicates genomes isolated from *S*. Typhi strain Ty2, STY indicates STY2879 gene fragment), and at different PC concentrations. Reaction colors were scored with the unaided eye as either pink, indicating no amplification; orange, indicating intermediate levels of amplification; or yellow, indicating full amplification until all the reactions reached a terminal yellow endpoint (Figure 2). Color outcomes shown in the table represent the median result across replicate reactions for each condition tested. All replicate reactions within each condition produced consistent amplification with no variation.

Primer set HJS-13-6 was then further tested with the more authentic positive control of genomic DNA from *Salmonella enterica* serovar Typhi strain Ty2 as the PC at a concentration of 10^3^ copies of DNA/µL. After no amplification was observed in 8% and 4% diluted whole milk (data not shown), amplification was observed in a 2% diluted raw milk matrix with a DTT of 30 minutes (Table 4).

## 4. Discussion and Conclusions

Due to the NEB colorimetric LAMP system’s potential for use in low-resource settings, we screened 17 primer sets targeting *S*. Typhi for their performance in this system. Fifteen novel primer sets were designed to target the segment of the STY2879 gene of *S*. Typhi originally identified by Fan et al. [12] to be specific to *S*. Typhi and highly conserved across *S*. Typhi isolates.

Fan et al. [12] established the STY2879 gene through multi-step comparative genomics and experimental validation using DNA from 22 geographically and temporally diverse *S*. Typhi isolates and 75 non-typhoidal *Salmonella* strains. From the process, the STY2879 gene emerged as a locus showing 100% positivity in all tested *S*. Typhi isolates and zero cross-reactivity with non-Typhi strains.

Subsequent independent clinical studies have further validated the diagnostic reliability of this target. Kaur et al. [13] incorporated STY2879-targeted primers into their magnetic nanoparticle–based “Miod” diagnostic platform for *S*. Typhi detection and observed no cross-reactivity with common bloodstream pathogens such as *Escherichia coli, Staphylococcus aureus, Pseudomonas aeruginosa, Acinetobacter baumannii, Enterococcus faecalis, Salmonella Paratyphi A*, and *Klebsiella pneumonia* at concentrations as high as 10^6^ CFU/mL in 28 clinical blood samples. In a more recent clinical study, Heamchandsaravanan et al. [14] confirmed the diagnostic performance of the STY2879 gene across 107 clinical blood samples.

To further support these prior findings, we conducted an *in silico* comparative analysis to assess the conservation of the STY2879 target gene segment used in this study. The inner primer regions (F2 and B2) are the first sequences to hybridize and initiate strand displacement synthesis. They define the core LAMP amplification target and are considered the key determinant of reaction specificity and efficiency [6]. Accordingly, the F2-B2 region was selected for alignment and comparative analysis. A BLASTN search using the F2-B2 sequence was performed against complete genome assemblies of *S*. Typhi (taxid: 90370). All 172 available complete *S*. Typhi genome assemblies exhibited 100% sequence identity to the F2-B2 region, indicating conservation of the LAMP amplification core across all sequenced *S*. Typhi strains (Table S3).

Collectively, the combination of previous experimental validation, independent clinical studies [12–14], and our *in silico* analysis demonstrates that STY2879 is a highly conserved and *S*. Typhi-specific target. The conservation of the F2-B2 inner primer region across all available *S*. Typhi genome assemblies supports its suitability for LAMP-based detection, reinforcing the robustness of STY2879 gene segment used in this study as a robust molecular target for rapid and accurate detection of *S*. Typhi with the HJS-13-6 assay.

Since LAMP primer sets can be hampered by non-specific amplification, the 15 novel primer sets and the primers from Fan et al. [12] and Abdullah et al. [17] were initially screened for the duration before nonspecific amplification occurred in the NEB system when no positive control was added (Figure 3). Three of the new primer sets showed a longer delay to non-specific amplification of at least 60 minutes when compared to either Fan et al. [12] or Abdullah et al. [17]. In the NEB system, both Fan et al. [12] and Abdullah et al. [17] primer sets showed that all NC reactions amplified within 40 minutes. This discrepancy may be due to the lack of optimization of reaction components, such as primers or magnesium, with the NEB system.

The three longest NC amplifying primer sets were further screened with positive controls. We found set HJS-13-6 had the largest time window (DTT) between amplification. Using DTT as a comparative metric provides a quantitative and standardized measure of how rapidly each primer set amplified true target DNA relative to any background signal, enabling objective selection of the most effective primer set for detecting the STY2879 target gene. Primer sets exhibiting large DTT values were interpreted as indicating faster and more robust amplification relative to background signal and as both efficient (rapid PC amplification) and specific (delayed or absent NC amplification).

Primer set HJS-13-6 was the most effective in detecting the target gene STY2879 in a water matrix using the STY2879 gene as the PC at a concentration of 10^5^ copies of DNA/µL. HJS-13-6 features overlapping F3 and F2 primer sites, which enabled the target segment to be suitable for incorporating loop primers into the design. The two–base pair overlap occurs at the 5′ end of the F2 region that does not participate in primer extension. Because FIP initiation relies on the 3′ end of F2 and tolerates a long 5′ overhang, this overlap is not expected to significantly affect LAMP kinetics or specificity. Any potential negative impact appears to be offset by the performance enhancement provided by the addition of the loop primers [18]. This is consistent with the displacement properties of Bst polymerase used in LAMP [19].

HJS-13-6 was further characterized for the limit of detection and tolerance for direct addition of milk to the reaction. HJS-13-6 demonstrates the ability to continue detecting low concentrations of STY2879 even when the final reaction volume is 2% milk (Table 5). To the best of our knowledge, this is the first study to demonstrate a successful *Salmonella enterica* serovar Typhi LAMP assay carried out with raw milk in the reaction matrix at a concentration of 2% V/V. The sensitivity of HJS-13-6 in the NEB system is estimated to be 2000 STY2879 copies/reaction or 1000 Ty2 genomes/reaction with a 1% raw milk matrix. This crude limit of detection estimation is based on the user having at least 20 minutes to interpret the color output.

**Table 5.**
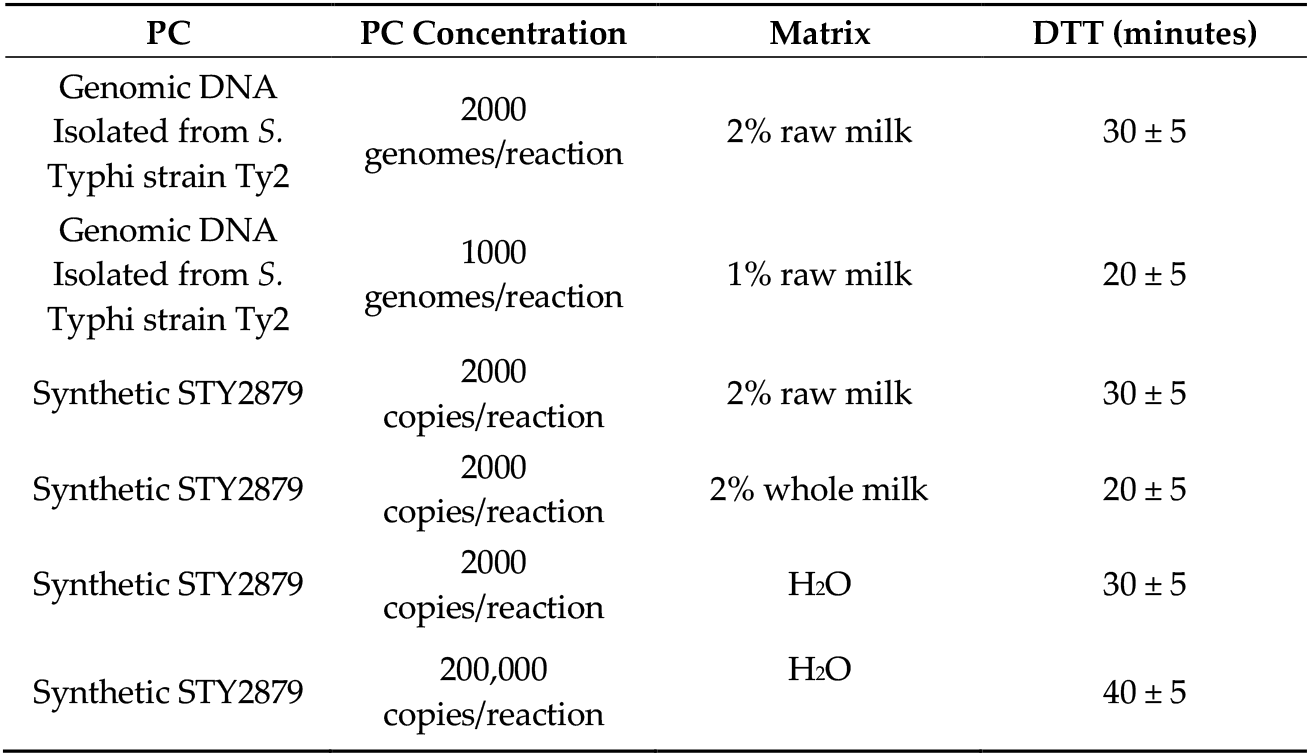
Effectiveness of the novel primer set HJS-13-6 across different matrices and varying sources and concentrations of STY2879, a previously validated sensitive and specific marker for *S*. Typhi. DTT values for each PC condition are shown. Larger DTT values indicate a better-performing assay.

| PC | PC Concentration | Matrix | DTT (minutes) |
| --- | --- | --- | --- |
| Genomic DNA<br>Isolated from *S.*<br>Typhi strain Ty2 | 2000<br>genomes/reaction | 2% raw milk | $30 \pm 5$ |
| Genomic DNA<br>Isolated from *S.*<br>Typhi strain Ty2 | 1000<br>genomes/reaction | 1% raw milk | $20 \pm 5$ |
| Synthetic STY2879 | 2000<br>copies/reaction | 2% raw milk | $30 \pm 5$ |
| Synthetic STY2879 | 2000<br>copies/reaction | 2% whole milk | $20 \pm 5$ |
| Synthetic STY2879 | 2000<br>copies/reaction | H<sub>2</sub>O | $30 \pm 5$ |
| Synthetic STY2879 | 200,000<br>copies/reaction | H<sub>2</sub>O | $40 \pm 5$ |

Fan et al. [12] and Abdullah et al. [17] report sensitivities of 15 copies/reaction and 20 CFU/reaction, respectively. HJS-13-6 in its current form is less sensitive than Fan et al. [12] or Abdullah et al. [17], but this limitation may be mitigated in three ways. First, the HJS-13-6 assay is not fully optimized with regard to reactant concentrations and reaction temperatures. Optimization can often lead to a 10-100 fold greater sensitivity [20]. Second, high assay sensitivity is not always critical for diagnostic test effectiveness, depending on the health thresholds and sample preparation steps. For example, Riyaz-Ul-Hassan et al. [21] found at least one raw dairy milk sample from India with a *Salmonella* concentration of ∼10^5^ cells/1 mL of milk. Additionally, samples are often concentrated or enriched before being moved into the LAMP reaction, thus increasing the initial *Salmonella* levels by orders of magnitude [12]. Finally, the sensitivity of HJS-13-6 is resistant to the input of milk (Table 5), indicating a robustness to any carryover of milk during the preprocessing. This robustness may allow for simplified sample processing and minimal wash steps after filtering, increasing the usability of the system in a low-resource setting in spite of the lower sensitivity.

The ability of HJS-13-6 to quickly detect STY2879 in various conditions (Table 5) warrants further studies of this primer system. Fan et al. [12] and additional studies [13,14] confirmed that STY2879 is a highly specific and conserved diagnostic target for *S*. Typhi. Future work should focus on improving the sensitivity of HJS-13-6 in the NEB system and on additional specificity tests of *Salmonella* strains and other common bacteria and pathogens that occur in raw milk, such as *Streptococcus thermophilus, Lactococcus lactis, Campylobacter* spp., *Escherichia coli*., *Listeria monocytogenes, Yersinia* spp., *Coxiella burnetii, Brucella* spp., and *Mycobacterium* spp. This study only included the addition of DNA directly to the assays and therefore does not address any complications that may come from a bacterial cell lysis or preamplification step. This critical part of the sample-to-answer process needs to be addressed in future research.

The novel HJS-13-6 primer set identified in this study is a first step in the development of a rapid *S*. Typhi test for raw milk samples prior to consumption. Due to the low resource demand of the colorimetric NEB LAMP system, HJS-13-6 has the potential to reduce the burden of global milk-borne outbreaks in raw milk-consuming communities in LICs.

## Data Availability

All data produced in the present study are available upon reasonable request to the authors.

## Supplementary Materials

Table S1: Sequence of the STY2879 target gene segment used in this study.; Table S2: Primer sequences of 15 novel LAMP primer sets designed for the STY2879 target gene fragment, reported in the 5’ to 3’ direction.; Table S3: BLASTN analysis of the F2-B2 region of the STY2879 target gene segment across complete *S*. Typhi genome assemblies, showing percent sequence identity for each strain.

## Author Contributions

Conceptualization, H.J.S. and T.E.R.; methodology, H.J.S. and T.E.R.; validation, H.J.S.; formal analysis, H.J.S.; investigation, H.J.S.; resources, H.J.S. and T.E.R.; writing— original draft preparation, H.J.S.; writing—review and editing, H.J.S. and T.E.R.; visualization, H.J.S.; supervision, T.E.R.; project administration, H.J.S.; funding acquisition, T.E.R. All authors have read and agreed to the published version of the manuscript.

## Acknowledgments

Significant contributions to the initial experimental designs, early data collection, and data analysis were made by Ansh Patel. This work was made possible thanks to generous financial support from Bob and Cathy O’Rear and the UT Austin College of Natural Sciences Freshman Research Initiative. We would like to acknowledge the support of the UT Austin Ellington Lab and the community of students that comprise the UT Austin FRI DIY Diagnostics Lab. The authors were gifted the enzymes used in this study from New England Biolabs in support of undergraduate research activities at the UT Austin.

## Abbreviations

The following abbreviations are used in this manuscript:

DTT: Differential time to threshold (time difference between negative control and positive control amplification)
LAMP: Loop-mediated isothermal amplification
F3: Forward Outer Primer
B3: Backward Outer Primer
FIP: Forward Inner Primer (composed of F1c and F2 regions)
BIP: Backward Inner Primer (composed of B1c and B2 regions)
LF: Loop Forward Primer
LB: Loop Backward Primer
PC: Positive control
NC: Non-template negative control

## References

1. Meiring, J.E.; Khanam, F.; Basnyat, B.; Charles, R.C.; Crump, J.A.; Debellut, F.; Holt, K.E.; Kariuki, S.; Mugisha, E.; Neuzil, K.M.; et al. Typhoid Fever. Nat. Rev. Dis. Primer 2023, 9, 71, doi:10.1038/s41572-023-00480-z.

2. Loor-Giler, A.; Sanchez-Castro, C.; Robayo-Chico, M.; Puga-Torres, B.; Santander-Parra, S.; Nuñez, L. High Contamination of Salmonella Spp. in Raw Milk in Ecuador: Molecular Identification of Salmonella Enterica Serovars Typhi, Paratyphi, Enteritidis and Typhimurium. Front. Sustain. Food Syst. 2025, 9, 1593266, doi:10.3389/fsufs.2025.1593266.

3. Grace, D.; Wu, F.; Havelaar, A.H. MILK Symposium Review: Foodborne Diseases from Milk and Milk Products in Developing Countries—Review of Causes and Health and Economic Implications. J. Dairy Sci. 2020, 103, 9715–9729, doi:10.3168/jds.2020-18323.

4. Nyokabi, S.; Luning, P.A.; De Boer, I.J.M.; Korir, L.; Muunda, E.; Bebe, B.O.; Lindahl, J.; Bett, B.; Oosting, S.J. Milk Quality and Hygiene: Knowledge, Attitudes and Practices of Smallholder Dairy Farmers in Central Kenya. Food Control 2021, 130, 108303, doi:10.1016/j.foodcont.2021.108303.

5. Amenu, K.; Wieland, B.; Szonyi, B.; Grace, D. Milk Handling Practices and Consumption Behavior among Borana Pastoralists in Southern Ethiopia. J. Health Popul. Nutr. 2019, 38, 6, doi:10.1186/s41043-019-0163-7.

6. Notomi, T. Loop-Mediated Isothermal Amplification of DNA. Nucleic Acids Res. 2000, 28, 63e–663, doi:10.1093/nar/28.12.e63.

7. Andrews, J.R.; Ryan, E.T. Diagnostics for Invasive Salmonella Infections: Current Challenges and Future Directions. Vaccine 2015, 33, C8–C15, doi:10.1016/j.vaccine.2015.02.030.

8. Snodgrass, R.; Gardner, A.; Semeere, A.; Kopparthy, V.L.; Duru, J.; Maurer, T.; Martin, J.; Cesarman, E.; Erickson, D. A Portable Device for Nucleic Acid Quantification Powered by Sunlight, a Flame or Electricity. Nat. Biomed. Eng. 2018, 2, 657–665, doi:10.1038/s41551-018-0286-y.

9. Chakraborty, S. Democratizing Nucleic Acid-Based Molecular Diagnostic Tests for Infectious Diseases at Resource-Limited Settings – from Point of Care to Extreme Point of Care. Sens. Diagn. 2024, 3, 536–561, doi:10.1039/D3SD00304C.

10. Jang, M.; Kim, S. Inhibition of Non-Specific Amplification in Loop-Mediated Isothermal Amplification via Tetramethylammonium Chloride. BioChip J. 2022, 16, 326–333, doi:10.1007/s13206-022-00070-3.

11. Jiang, Y.S.; Bhadra, S.; Li, B.; Wu, Y.R.; Milligan, J.N.; Ellington, A.D. Robust Strand Exchange Reactions for the Sequence-Specific, Real-Time Detection of Nucleic Acid Amplicons. Anal. Chem. 2015, 87, 3314–3320, doi:10.1021/ac504387c.

12. Fan, F.; Du, P.; Kan, B.; Yan, M. The Development and Evaluation of a Loop-Mediated Isothermal Amplification Method for the Rapid Detection of Salmonella Enterica Serovar Typhi. PLOS ONE 2015, 10, e0124507, doi:10.1371/journal.pone.0124507.

13. Kaur, A.; Kapil, A.; Elangovan, R.; Jha, S.; Kalyanasundaram, D. Highly-Sensitive Detection of Salmonella Typhi in Clinical Blood Samples by Magnetic Nanoparticle-Based Enrichment and in-Situ Measurement of Isothermal Amplification of Nucleic Acids. PLOS ONE 2018, 13, e0194817, doi:10.1371/journal.pone.0194817.

14. Heamchandsaravanan, A.R.; Shanmugam, K.; Perumal, D.; Shankar, D.; Kalpana, S.; Dhandapani, P. Loop-Mediated Isothermal Amplification (LAMP) Assay Targeting STY2879 Gene for Rapid Detection of Salmonella Enterica Serovar Typhi in Blood. Res. J. Pharm. Technol. 2024, 2087–2092, doi:10.52711/0974-360X.2024.00330.

15. New England Biolabs NEB LAMP Primer Design.

16. Colosimo, F.; Tanner, N.A.; Patton, G.C.; New England Biolabs LAMP Primer Design Using the NEB LAMP Primer Design Tool: Critical Considerations for Assay Robustness, Speed and Sensitivity; New England Biolabs, 2025;

17. Abdullah, J.; Saffie, N.; Sjasri, F.A.R.; Husin, A.; Abdul-Rahman, Z.; Ismail, A.; Aziah, I.; Mohamed, M. Rapid Detection of Salmonella Typhi by Loop-Mediated Isothermal Amplification (LAMP) Method. Braz. J. Microbiol. 2014, 45, 1385–1391, doi:10.1590/S1517-83822014000400032.

18. Nagamine, K.; Hase, T.; Notomi, T. Accelerated Reaction by Loop-Mediated Isothermal Amplification Using Loop Primers. Mol. Cell. Probes 2002, 16, 223–229, doi:10.1006/mcpr.2002.0415.

19. Oscorbin, I.; Filipenko, M. Bst Polymerase — a Humble Relative of Taq Polymerase. Comput. Struct. Biotechnol. J. 2023, 21, 4519– 4535, doi:10.1016/j.csbj.2023.09.008.

20. Wang, D.-G.; Brewster, J.; Paul, M.; Tomasula, P. Two Methods for Increased Specificity and Sensitivity in Loop-Mediated Isothermal Amplification. Molecules 2015, 20, 6048–6059, doi:10.3390/molecules20046048.

21. Riyaz-Ul-Hassan, S.; Verma, V.; Qazi, G.N. Real-Time PCR-Based Rapid and Culture-Independent Detection of Salmonella in Dairy Milk - Addressing Some Core Issues. Lett. Appl. Microbiol. 2013, 56, 275–282, doi:10.1111/lam.12046.

